# Responsibility for vitamin D supplementation of elderly care home residents in England: falling through the gap between medicine and food

**DOI:** 10.1101/2020.06.21.20136697

**Authors:** Joseph Williams, Carol Williams

**Affiliations:** Brighton and Sussex Medical School, Division of Medical Education - Postgraduate Medicine, University of Brighton, Watson Building, Falmer, BN1 9PH; Health Promotion & Public Health, School of Health Sciences, University of Brighton, Westlain House, Falmer, BN1 9PH

## Abstract

**Introduction:** Vitamin D supplements are recommended for elderly care home residents with little sunlight exposure. However, their use in care homes is limited and vitamin D deficiency in residents is widespread. This study aimed to understand perceived responsibility for the vitamin D status of residents and the determinants of current practice around supplementation.

**Methods:** Thirteen semi-structured interviews were conducted with key informants in two areas of Southern England including care home managers, general practitioners (GPs) and public health professionals. Interviews were audio recorded and transcribed verbatim.

**Results:** Inductive thematic analysis identified four themes – medical framing; professional and sector boundaries; awareness of national guidance; and ethical and practical issues. Vitamin D supplements were not routinely given to residents, and most participants, including the GPs, believed the vitamin D status of residents was the responsibility of the GP. Care home managers felt unable to make decisions about vitamin D and vulnerable to suggestions of wrongdoing in administering over-the-counter vitamin tablets. This results in vitamin D requiring prescription by medical professionals and few care home residents receiving vitamin D supplements.

**Conclusion:** The medical framing of vitamin D supplements in care homes is a practical barrier to residents receiving them and is out of step with public health recommendations. Vitamin D levels in care home residents could be improved through universal supplementation. This requires a paradigm shift so that vitamin D is understood as a protective nutrient as well as a medicine, and a public health as well as a medical responsibility. The failure to ensure vitamin D adequacy of residents may emerge as a factor in the spread and severity of COVID-19 in care homes and gives increased urgency to addressing this issue.

**KEY MESSAGES:** *What is already known about the subject?:* Nutrition guidelines recommend elderly care home residents take vitamin D supplements as a preventative measure. This is rarely implemented in practice and vitamin D inadequacy is widespread.

*What are the new findings?:* Medical framing of vitamin D in the care sector puts elderly residents at risk of vitamin D deficiency. Vitamin D supplements are perceived as medicines requiring an individual prescription and diagnosis by a medical professional. This is out of step with public health recommendations. The system’s failure to protect the vitamin D status of the elderly in care homes may have implications in the context of COVID-19.

*How might these results change the focus of research or practice?:* Prompt a review of current guidelines and regulations in England to establish responsibility for implementing public health recommendations on vitamin D supplementation in care homes. Further research on feasibility of implementation strategies is needed.

## INTRODUCTION

Public health is classically defined as “the art and science of preventing disease, prolonging life and promoting health through the organized efforts of society”.^(1)^ Arguably societies organise themselves more readily to improve the health of those in view. Those out of sight, including older adults in residential care settings, can often be forgotten. The coronavirus (COVID-19) pandemic has brought the health and vulnerability of those in elderly care homes to the fore, yet some feel action to tackle to contain the virus in social care has been late and inadequate.^(2, 3)^ A. possible link between low vitamin D status and severity of COVID-19 is emerging.^(4, 5)^ Our investigation into how vitamin D nutrition supplementation is addressed in care homes reveals both a failure to implement evidence-based recommendations and a social injustice in urgent need of public health advocacy and resolution.

Vitamin D is a prohormone rather than a nutrient, and the main source is endogenous synthesis in the skin rather than ingestion from food. Synthesis occurs when skin is exposed to ultraviolet B (UVB) radiation in sunlight. In winter months at latitudes with shorter day lengths, the UVB is insufficient due to the low angle of the sun, thus vitamin D deficiency can be viewed as sunshine deficiency.^(5, 6)^ There are few significant food sources and it is usually not possible to meet vitamin D needs from diet alone.^(7)^

Vitamin D is required for the regulation of calcium and phosphorus metabolism and there is strong evidence that insufficient vitamin D affects musculoskeletal health and development.^(8)^ Vitamin D has also been cited as having a potential role in numerous other aspects of health, including immunity, cardiovascular health, neurological conditions, respiratory infections, lung function and cancer.^(8, 9)^ There has been recent interest in the potential role of vitamin D deficiency in the severity of COVID-19,^(10)^ and possibly its contagion.^(4)^ COVID-19 mortality has been higher in older age groups and Black, Asian and minority ethnic groups.^(4, 11)^ These groups are at increased risk of not getting sun exposure for vitamin D synthesis due to spending less time outside, covering their skin, or having darker skin which needs longer exposure for synthesis to occur.^(12, 13)^

Elderly residents in residential care homes / nursing homes (hereafter referred to as care homes), particularly those with limited mobility, are likely to spend more time indoors and have limited sun exposure. They have been recognised as a group vulnerable to vitamin D deficiency and requiring supplements for nearly 30 years.^(8, 14, 15)^National bodies from countries around the world have issued similar recommendations for care home residents throughout this time period, including Australia, Canada, France, Norway, New Zealand, USA.^(16, 17)^

Despite the decades of recommendations for supplementation in this population, vitamin D deficiency in care home residents is widespread throughout Europe, Asia and Americas.^(16)^ Studies in Austria,^(18)^ Belgium,^(19)^ Germany,^(20)^ and Sweden^(21)^ found that almost all residents of care homes were vitamin D deficient. A notable exception is New Zealand where a publicly funded universal vitamin D supplementation programme for care homes has been in operation since 2011. An evaluation in 2014 found that 75% of care home residents took supplements, and almost all of those receiving the supplements had healthy serum levels of 25-hydroxyvitamin D (25(OH)D), the marker for vitamin D status.^(22)^

Whether vitamin supplements are considered a medicine, or a food has significant implications. In most countries vitamin and mineral supplements are regulated and sold as foodstuffs rather than medicines.^(23-25)^ Supplementation with vitamin D as a food is considered a personal responsibility which, in a care home setting, may be passed on to family members or staff. However, if vitamin D supplements are considered to be medicines, responsibility is deferred to medical professionals. The duality of vitamin D as both medicine and food is evident in contradictory health and care regulations and guidance in England. Care homes are required to assess resident’s the nutritional needs and provide food to meet those needs, including “dietary supplements when prescribed by a health care professional”.^(26)^ Guidance to NHS clinical commissioning groups expressly advises GPs against routine prescription of vitamins in primary care due to “limited evidence of clinical effectiveness” and because “vitamin D supplements can be bought cheaply and easily”.^(27)^ The guidance makes exceptions for medically diagnosed vitamin D deficiency or for osteoporosis but not for maintenance or preventative treatment. The purpose of this study was to better understand current practice in the implementation of public health guidance on vitamin D supplementation in the care home setting.

## METHODS

The research was carried out using a pragmatic interpretive methodology. Semi structured interviews were used to explore current practice around vitamin D supplementation, perceived responsibility for vitamin D status and supplementation, and barriers to supplementation. Interviews were conducted by telephone with key stakeholders identified as having a role in the care of elderly care home residents. The study used purposive opportunistic sampling to recruit participants, including senior members of care home staff; general practitioners (GPs); members of local authority public health departments; and other relevant professionals (as identified by participants) with shared constituencies of care home residents. Eligible participants were approached by email and invited to participate. Informed consent was obtained from all participants before commencing the interview. Recruitment was until data saturation was reached.

### Data collection and analysis

Interviews were conducted between 22 April 2018 and 31 August 2018, audio recorded and transcribed verbatim. The data was approached inductively using a six-stage process as described by Braun & Clarke.^(28)^ JW read and re-read the transcripts for immersion in the data and developed initial codes and themes. All transcripts were independently read and coded by CW, using qualitative data software (NVivo) to support the process. Codes and themes were elaborated using an iterative process, with the two researchers referring-back to the raw data to substantiate emerging ideas.

## RESULTS

Thirteen interviews, lasting approximately 15 minutes, were conducted with employees of 13 different organisations operating in two districts of non-neighbouring areas of South East England. Participant roles and sites are outlined in Table 1. Four broad themes and six sub themes were identified, these are summarised in Table 2 and described in detail below.

**Table 1.** Participant demographics.

| Areas | Coding | Position and organisation | Organisation |
| --- | --- | --- | --- |
| A | GP1, GP2 | 2 general practitioners | 2 general practices |
|  | CHM1, CHM2 | Two care home managers | Two care homes |
|  | PubH1 | Senior public health practitioner | Local authority |
| B | GP3, GP4 | Two general practitioners | Two separate general practices in this area |
|  | CHM3, CHM4 | Two care home managers | Two care homes |
|  | FallSP | Falls specialist practitioner | NHS community trust |
|  | EConsP | Consultant elderly care physician | NHS hospital trust |
|  | PCDiet | Primary care dietitian | NHS clinical commissioning group |
|  | PubH2 | Public health programme manager | Local authority |

**Table 2:** Summary of themes and subthemes.

|  |
| --- |
| Themes and sub-themes |
| 1. Medical framing: Vitamin D understood as a medical issue |
| 2. Professional and sector boundaries<br>○ Perceived responsibility and roles: authority and vulnerability<br>○ Cost implications<br>○ Policy, regulations, guidance and compliance |
| 3. Low awareness of recommendations and need for vitamin D supplements. |
| 4. Ethical and practical considerations<br>○ Ethics of consent and life stage<br>○ Resistance to polypharmacy |

**Table 3:** Representative quotations for Theme 1: Vitamin D understood as medical issue.

| Theme 1: Vitamin D understood as medical issue |
| --- |
| *“Quite a lot of the vitamin D stuff that we tend to prescribe is mixed in with calcium as well... Usually that will be in people who are either known to be osteoporotic or may have had a previous low trauma fracture before.”* (GP1)<br>*“We don't have anything in place in terms of a protocol for specifically checking it with each patient. It tends to come up in those where it's more obvious because of their history. And if we happened to think about it, which probably means missing quite a few patients who benefit from it.”* (GP2)<br>*“No. Literally since I've been here, there's only been two occasions where residents have gone on to vitamin D. And that's because they've had routine blood tests done, and it's identified their levels were low.”* (CHM4) |

**Table 4:**
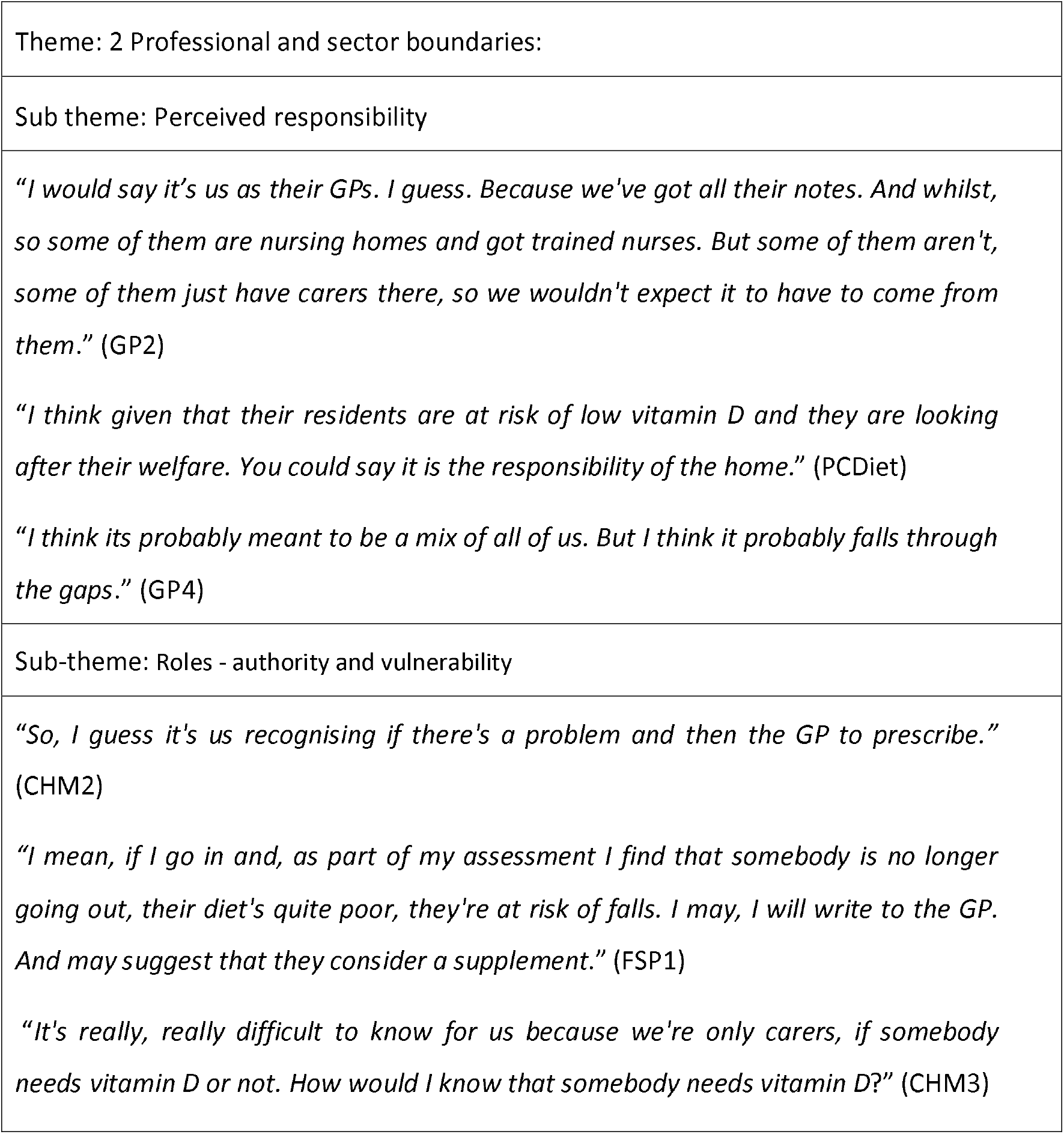
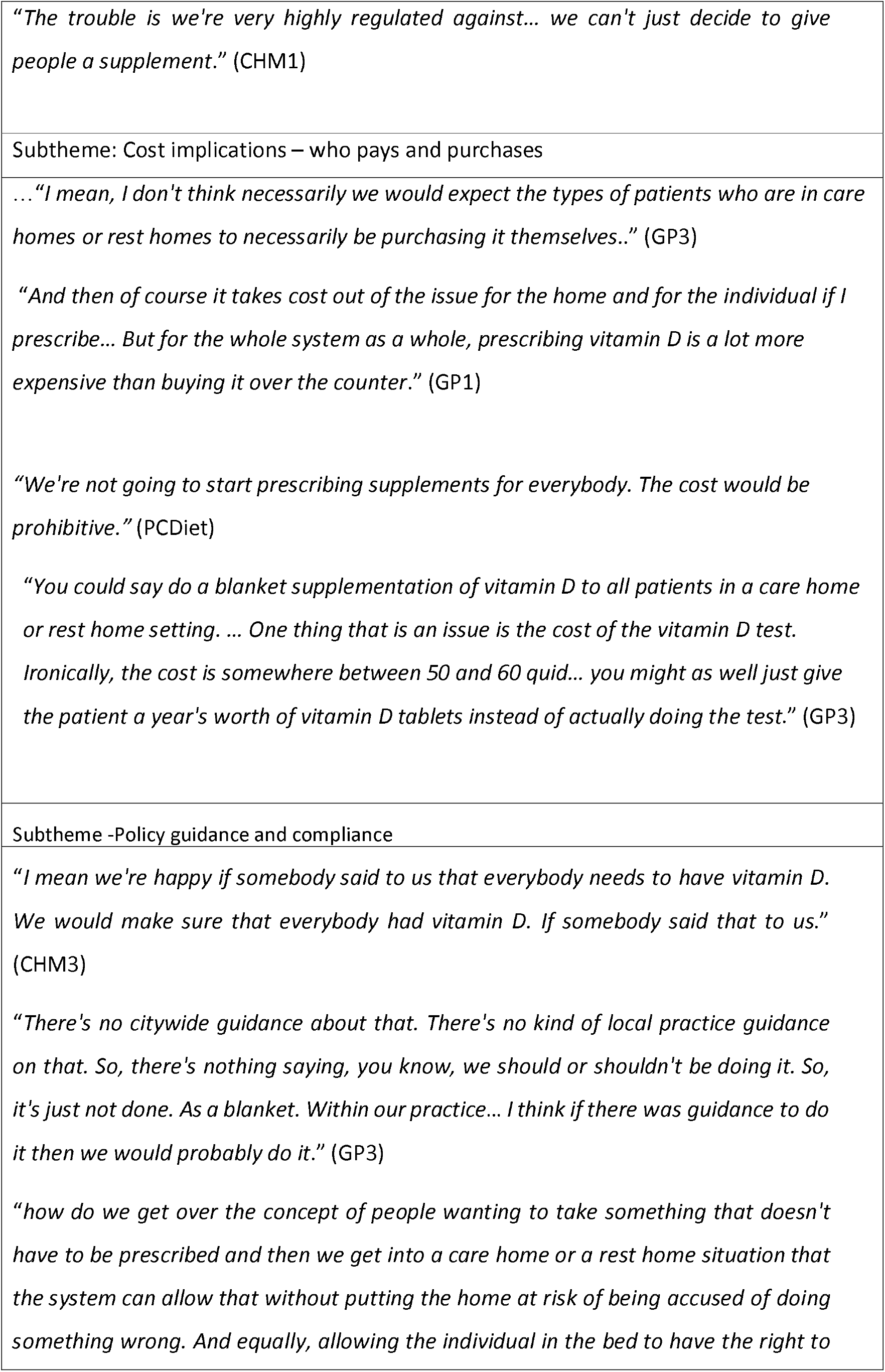
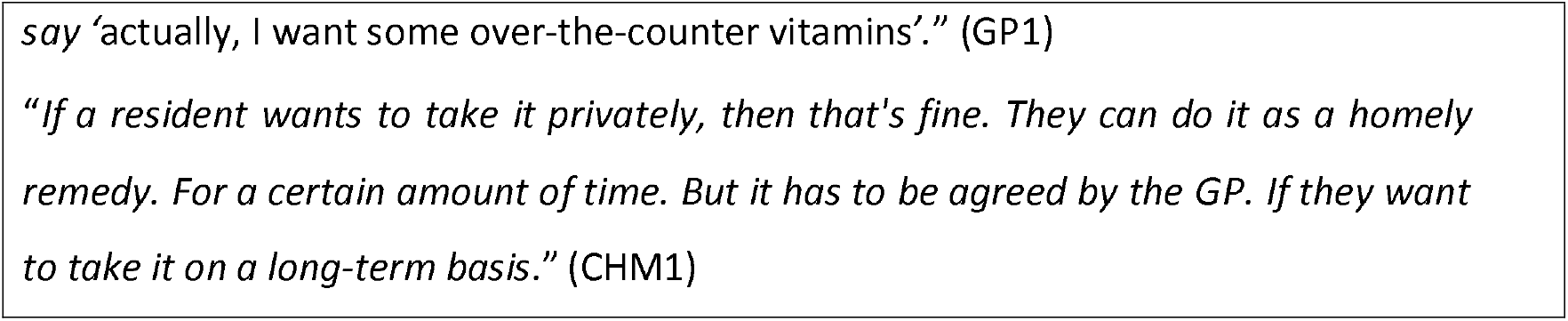
Representative quotations for Theme 2: Professional and sector boundaries.

**Table 5:**
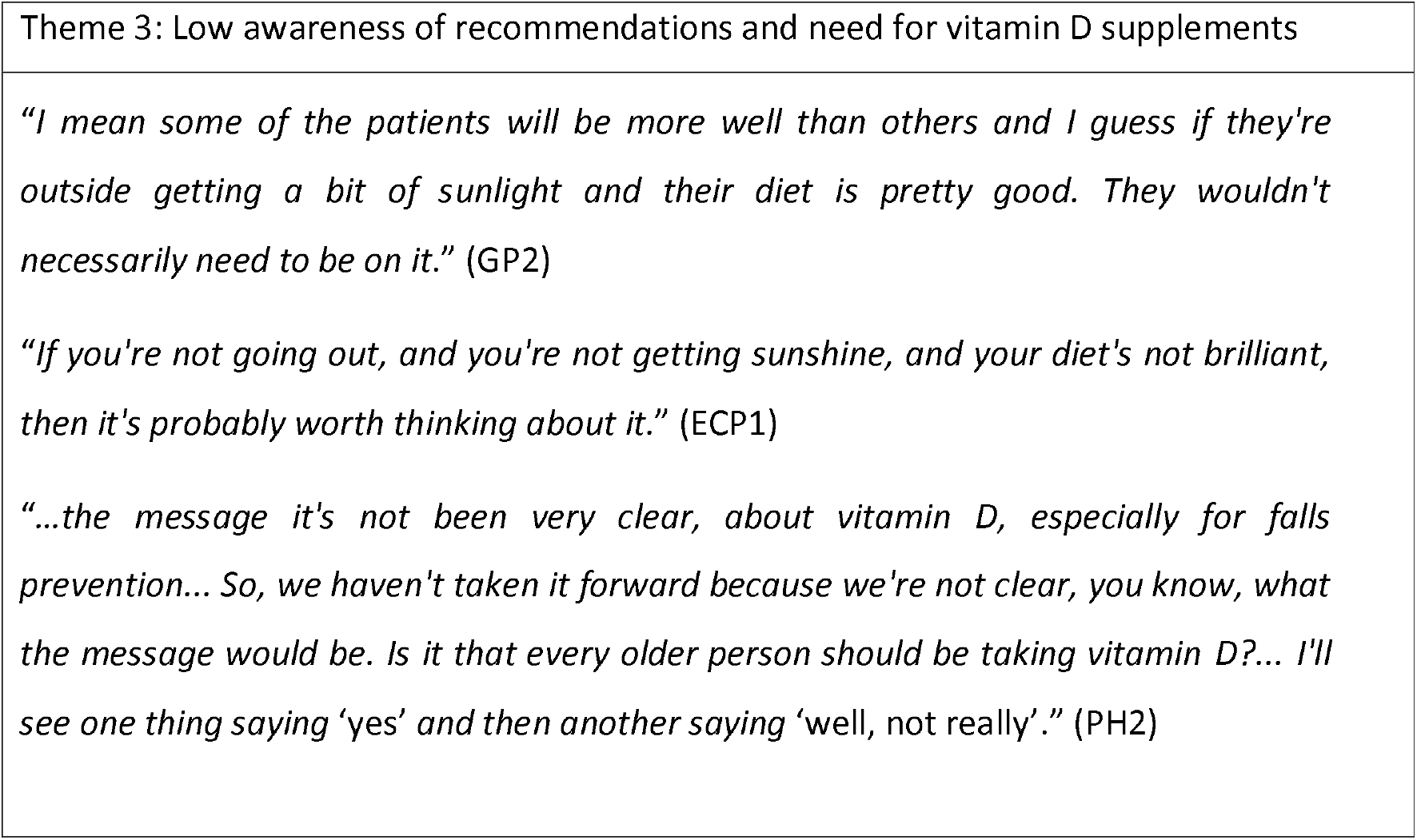
Representative quotations for Theme 3: Low awareness of recommendations and need for vitamin D supplements.

**Table 6:**
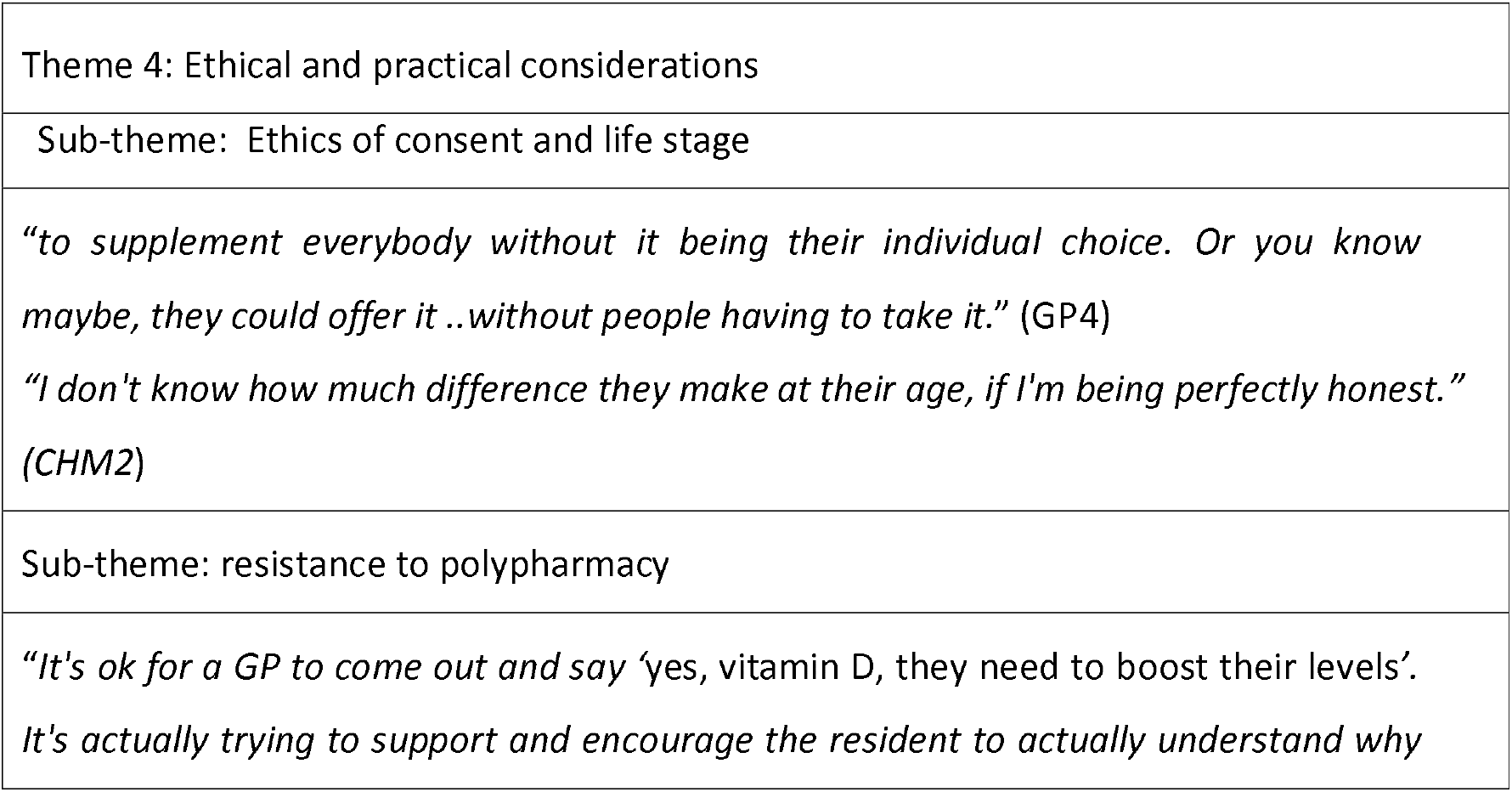
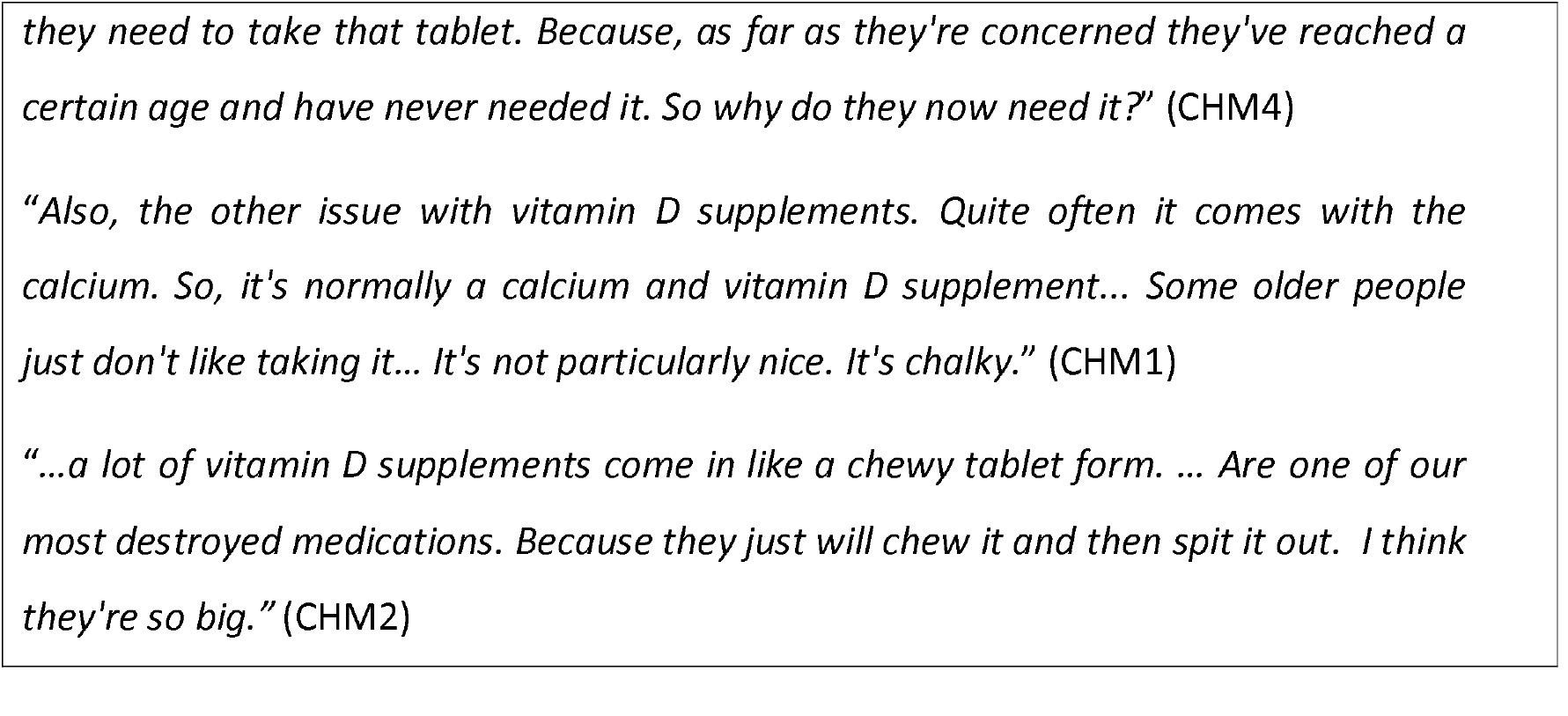
Representative quotations for Theme 4: Ethical and practical considerations.

### 1. Vitamin D understood as a medical issue

All the stakeholders in elderly care we spoke to referred to vitamin D as something which residents took on an individual basis, usually because it had been prescribed by a GP. None knew of any care home where vitamin D supplements were given routinely to residents as part of a protective public health measure; ‘*So there’s no kind of protocol or carpet / universal plan to prescribe vitamin D supplements to patients of ours at care homes*’ (GP3). GP prescribing was contingent on a formal diagnosis of vitamin D deficiency, arthritis, or osteoporosis or following falls or fractures.

### 2. Professional and sector boundaries

#### Perceived responsibility and roles

There was a strong agreement, including from the GP participants, that the vitamin D status of elderly care home residents was the responsibility of the GP. Only the dietitian suggested perhaps the primary responsibility should lie with the care home since the home had a “*responsibility to provide a diet that includes all the nutrients they need*.*”* PCDiet. One of the GPs touched on the issue of medicine versus diet that is central to this subject “*…if you’re looking at it from a medical point of view, then I, the GP, would be responsible. If you’re looking at this from a ‘are the patients getting a well-balanced diet that would include vitamin D… … I would see that as the home’s responsibility*.” (GP1). Several of the non-medical participants felt the responsibility was shared between the care home and the GP, but here the GP was the decision maker and their role was to alert GPs about individual residents. From the interviews it was clear that there was an implicit hierarchy and deferment to the medical profession which meant care home staff felt they would not be able to make decisions about supplements: “*we can only ask the doctor if they will prescribe it*.” (CHM1). Respondents from the medical profession acknowledged and understood why care home staff may feel constrained in this area.

#### Cost implications – who pays and purchases?

There was considerable ambiguity about who should be responsible for providing and paying for supplements. Again, most participants felt this was a GP responsibility because of their role as prescriber, and any change from this arrangement would have financial consequences for the care home or the residents. The theoretical responsibility of individual residents paying for their own over-the-counter (OTC) supplements and the impractical reality of this for residents with mobility or cognitive constraints was acknowledged. One GP participant considered the financial aspects more broadly contrasting the cost of testing serum vitamin D levels which is far higher than the cost of a year’s supply of vitamin D supplements.

#### Policy guidance and compliance

Each implementing stakeholder we spoke to was adhering to the regulations and operating guidance in their sector, but the absence of any specific guidance about how to implement public health advice on vitamin D supplementation, these restricted homes to only dispensing vitamin D on prescription: “*If somebody said to us that everybody needs to have vitamin D we would make sure that everybody had vitamin* D” (CHM3). ‘Homely Remedy’ policies can provide a mechanism for homes to give over-the-counter products to residents,^(29)^ and three of the four care homes had a homely remedy policy in place. However, they explained that the GP needs to be involved for the supplementation to continue in the long-term.

### 3. Low awareness of national guidance and need for vitamin D supplements

Except for the dietitian, none of the participants seemed familiar with the recommendation that all elderly care home residents should receive vitamin D without the need for pre-assessment: *“How would I know that somebody needs vitamin D*?” (CHM3). Some participants highlighted the role of diet and sunlight in vitamin D status and seemed unaware that these sources cannot provide adequate levels even for the general population during winter months. Several of the GPs referred to care homes needing to ensure that residents got out into the sun, which could be effective in the summer months, but mobility issues mean this is impractical for many residents.

### 4. Ethical and practical considerations

When asked about universal supplementation, several participants raised ethical issues, both around the population versus individual approach and the limited ability of some residents to give consent: “*We have a lot of people with dementia who can’t make that decision for themselves*” (CHM1). Others questioned the value and ethics of introducing another tablet given the age of residents

#### Resistance to polypharmacy

Care home staff are responsible for the administration of medicines and supplements to residents and highlighted practical issues which would be barriers to wider supplement use. This included the number of medicines residents already take and how introducing more tablets would add to the pill burden for both residents and staff. Another common point related to the formulation of vitamin D with calcium and how it was difficult to take. Crucially, these are not generic medication compliance issues since in tablet form, even high-strength vitamin D tablets are very small.

## DISCUSSION

This study aimed to understand perceived responsibility for the vitamin D status of care home residents and explore factors influencing supplementation practice. The study found an overwhelmingly medical rather than public health conception of vitamin D by those involved in the welfare of care home residents. This stems from regulatory and professional boundaries which treat vitamin D as a medicine and deny residents recommended dietary supplements to protect their vitamin D levels. These findings may be used to inform advocacy for policy and practice change in England and other countries where vitamin D supplementation in elderly care homes tends to be by prescription-only.

### Medicine or food

The participants of this study all saw vitamin D supplementation as the responsibility of the GP. This medical framing means care home staff fear overstepping their role and defer to medical professionals to diagnose vitamin D deficiency or osteoporosis as a medical condition then prescribe vitamin D as a “medicine” in response. In this respect, both care home staff and GPs are complying with the respective regulations and guidance governing their sector: care home staff to administer supplements only when they are prescribed; and GPs not to prescribe supplements for prevention or maintenance. As a result, few elderly care home residents receive vitamin D supplements.

The medicalisation of this nutrient is at odds with the UK’s nutrition advisory committee Scientific Advisory Committee on Nutrition (SACN), and other nutrition expert panels, who consider vitamin D supplements, along with natural food sources and fortified foods, as a dietary source.^(8)^ It is also out of step with the public health recommendations which identify a population-wide need for vitamin D. Low awareness of the public health need for supplements is not unique to the care sector, other reports have noted that many health professionals may be not be aware of issues around vitamin D including synthesis, diet, and the importance of supplements.^(30)^ Furthermore, national guidance on healthy catering for food served to older people in residential care makes a general statement on the need for vitamin D supplements without addressing how this happens in practice or who is responsible.^(31)^

Our findings suggest that in the elderly residential care sector, the narrative of vitamin D as a treatment is at the expense of any parallel narrative of vitamin D as a protective and preventative public health measure. Regulations and governance protocols must ensure both approaches are permitted. As agencies review evidence on vitamin D supplements and COVID-19 there is an opportunity to address the practice gap in care homes by including specific mechanisms for delivery.^(32)^ In time, a reliable preventative supplementation strategy should lessen the need for treatment of deficiency.

### Social justice perspective

The recommendations for vitamin D rely on personal responsibility and it is questionable whether this is appropriate in populations with limited autonomy. Community dwelling elderly and independent care home residents can purchase their own vitamin D supplements for personal use. However, people usually move into care homes because they no longer have mobility or mental capacity to live independently, and accordingly have more limited control over lifestyle decisions. Most elderly care home residents have some form of dementia.^(33, 34)^ This results in a two-tier system, discriminating against those with least cognitive independence. Even if residents or their family purchase OTC vitamin D supplements and request that these are administered by care staff under a homely remedy policy, the GP still needs to approve their use. Homely remedies are defined as OTC products for short term treatment of minor ailments, so do not encompass daily vitamin supplements.^(29, 35)^ The regulatory environment care homes operate in would make it difficult for a care home to unilaterally decide to offer all residents daily vitamin D supplements, not least because residents in care homes are frequently registered with different GPs.

### Finance

Even in a universal healthcare system such as the National Health Service, there are significant and often competing financial considerations for the different agencies involved in elderly care. The care home sector in England is fragmented with more than 15,000 residential and nursing care homes operated by more than 5,000 providers.^(36, 37)^ Funding for places comes from local authorities, NHS, charities, or directly from residents and their families. As highlighted by one of the participants, the involvement of GPs means care homes do not need to purchase supplements. However, GP time is scarce and costly and using NHS prescribing as a vehicle for getting vitamin supplements to a sizable at-risk population is a poor use of NHS resources. One year’s supply of OTC vitamin D costs approximately £15.00 per person at typical high street prices. These costs could be absorbed into existing care home fees or covered by local authorities. To ensure that children under five years receive vitamin D, low income families on benefits are eligible for free NHS Healthy Start vitamin drops and some local authorities fund Healthy Start vitamins for all children.^(38)^ We believe equivalent arrangements should be in place for frail elderly.

### Formulation of vitamin D

Preventative dose vitamin D supplements can be formulated as tiny tablets, and are also available to be administered as drops, chewable and dispersible formulations, thus is easier to take than when combined with calcium. However, the additional burden to care staff of being required to administer a supplement in any form needs to be acknowledged. Provisioning vitamin D through an appropriately fortified food stuff would shift the delivery firmly to care homes as part of their food service. This would also eliminate the need for revision of existing controls on medicines in care homes. However, the literature on use of vitamin D fortified foods in the elderly is limited and the feasibility needs further research. ^(39, 40)^

### COVID-19 and the ethics of inaction

Many frail elderly in the community suffer undernourishment but the move into a care home can improve nutrition as meals are provided and residents receive support with eating. Vitamin D needs to be part of this care. Other commentators in this journal have noted that policies and recommendations on vitamin D do not seem to be “taken seriously enough”.^(4)^ Low vitamin D status exposes the elderly to a wide range of increased risks, including falls,^(8)^ seasonal flu and possibly COVID-19.^(9)^ Whatever the outcome of research into the links between COVID-19 and vitamin D, the pandemic has brought conditions in care homes into the public eye and on to the political agenda. Whilst practices in care homes are in the spotlight there is an urgent need to for action to ensure vitamin D recommendations can be applied in care homes. If it proves helpful in preventing further COVID-19 deaths, we can be glad we acted.

### Limitations of the study

While this study provides insight into some of the underlying reasons why care home residents tend not to receive preventative vitamin D supplements, it is a small study and practice may be different in other areas. We used purposive sampling in two non-neighbouring areas of southern England, and included stakeholders employed by a range of organisations to improve the generalisability. Stakeholders reference to national policy drivers as key determinants of current practice suggests the finding may be generalisable more widely in England. The literature suggests that the core issue of medicalisation of vitamin D may be relevant across the care sector in other countries.^(16)^

## CONCLUSION

The general population of the UK are recommended to take a daily vitamin D supplement.^(8)^ Residents of elderly care homes are particularly at risk but have limited ability to make lifestyle decisions of this kind. This study highlights that a gap exists between guidance and practice around vitamin D supplementation in this population. Professionals involved in the care of elderly residential care home residents perceive vitamin D as a medicine rather than a food. This means not enough elderly care home residents receive vitamin D supplements.

As a result, their vitamin D levels remain low and they are at increased risk of falls and fractures and possibly greater severity of COVID-19.

Vitamin D in care homes needs further attention with mechanisms developed for the administration of low-risk dietary supplements or fortified foods by care staff. This will help us to progress from a situation where care staff feel constrained and vulnerable, to one where they are supported to improve the health of those in their care. For a universal, population-based approach, vitamin D supplementation at protective levels needs a professional separation from medicine and reframing as a matter of public health nutrition.

## Data Availability

Data not publicly available because interview transcripts contain information that could compromise research participant privacy and consent. Data redacted to ensure anonymity are available to researchers from the corresponding author upon reasonable request.

